# Antibiotic Seeking Pathways and Patterns of Usage among Patients with Productive Coughs Attending Selected Chest Clinics in Nairobi County, Kenya

**DOI:** 10.64898/2026.04.04.26350147

**Authors:** Alvin Kinji Mwabu, Winnie C. Mutai, John Kiiru, John Mwaniki, Walter Jaoko

## Abstract

**Introduction:** Antibiotic misuse is a major driver of antimicrobial resistance (AMR), contributing to an estimated 1.27 million deaths globally. In Kenya, inappropriate antibiotic use is shaped by health-seeking behaviors and sociodemographic factors. However, little is known about how adults with productive coughs seek and use antibiotics, or how sociodemographic factors underpin these practices. This study explored antibiotic-seeking pathways, usage patterns, and the sociodemographic factors influencing these practices among adults with productive coughs attending selected chest and tuberculosis clinics in Nairobi County, Kenya.

**Methodology:** A facility-based cross-sectional study was conducted among 400 adults (≥18 years) with productive coughs. Data were collected using a structured questionnaire on sociodemographic characteristics, antibiotic-seeking pathways, and use patterns.

**Results:** Most participants were male (65.0%) and employed (67.0%), with 68.3% earning below Ksh 10,000 (approximately USD 80) monthly and 35.8% having basic education. A history of smoking (37.3%), tuberculosis (32.0%), or other comorbidities (29.8%) was common. Among 347 (86.7%) antibiotic users, 46.4% obtained antibiotics through general practitioners (GP) only, 31.4% via both GP and over-the-counter (OTC) sources, 15.3% from OTC only, and 6.9% through self-medication. Females were more likely to self-medicate (13.3% vs. 3.2%) and had higher odds of antibiotic use (cOR: 2.00; 95% CI: 1.04–4.10). Tuberculosis history was linked to greater GP reliance (61.7% vs. 37.4%). Low-income participants mainly used GP-only sources, while higher-income earners favored GP plus OTC routes (RRR: 2.67; 95% CI: 1.41–5.05). Empirical use was common (71.1%), dominated by Amoxicillin (90.8%), with multiple antibiotic use reported by 67.2% of the participants.

**Conclusion:** Antibiotic use among adults with productive coughs in Nairobi was widespread and largely empirical, dominated by Amoxicillin and Amoxicillin/Clavulanic acid. Self-medication, unregulated antibiotic access, and inappropriate use highlight the urgent need for stricter prescription enforcement and strengthened stewardship programs to promote rational antibiotic use and curb AMR.

## INTRODUCTION

Respiratory diseases affect over a billion people globally (1), with rhinoviruses responsible for 30–50% of cases, coronavirus for 10–15%, and 30–40% having unknown causes (2). Although many respiratory infections are self-limiting, they remain a significant driver of antibiotic misuse, which contributes to the rise in antibiotic resistance (ABR). Among these illnesses, cough is a common symptom that plays a role in clearing the airways (3), but persistent coughs can negatively impact quality of life (4), prompting many patients to seek antibiotics.

Among the most common antibiotic seeking pathways is consulting general practitioners (GP). Cough-related GP visits exceed 30 million annually (5, 6). Respiratory tract infections (RTIs) account for 20% to 90% of all prescriptions in Europe (7), with 60% of these occurring in the UK alone (8). In China, over 50% of RTI patients received antibiotics, 78% for colds and 93% for acute bronchitis (9). In developing nations, antibiotics were prescribed for over 80% of RTIs in Kenya and 77.6% in Uganda respectively (10, 11).

However, this data is largely provider-driven and post-diagnostic, reflecting only a portion of the antibiotic-seeking landscape. The ’symptom iceberg’ phenomenon (12) illustrates that many individuals self-treat symptoms like cough without engaging formal healthcare. A significant number of patients obtain antibiotics over the counter (OTC), bypassing prescription requirements. Despite legal restrictions (13), over 50% of antibiotics worldwide are obtained without prescriptions (14), a trend particularly evident in developing countries (15).

In China, inadequate antibiotic stewardship in pharmacy contribute to an “access-excess” dilemma (16), and in some low- and middle-Income countries (LMICs), 100% of pharmacies dispense antibiotics OTC for upper respiratory tract infections (URTIs) (17). According to (18), RTIs patients often obtain antibiotics from pharmacies (65.5 %), and drug shops (37.5 %). Additionally, patients also self-medicate using leftover antibiotics or medications shared by others in case they do not seek formal healthcare.

Symptoms such as sore throat, swollen tonsils and redness driver such practices in Europe (19, 20). These informal pathways of acquiring antibiotics, whether through pharmacies, drug stores, social networks and rural vendors (18), often result into misuse. Misdiagnosis, inappropriate drug choices, delayed treatment, and drug interactions due to self-medication contribute ABR (21, 22). Additionally, improper prescribing behaviors like overuse, underuse or incorrect drug regimens for RTIs further increase ABR risks (23).

In Kenya, widespread RTIs and both formal and informal access to antibiotics create favorable environment for misuse. Exposure to antibiotics increases the likelihood of colonization by resistant bacterial pathogens linked to 4.95 million deaths globally in 2019 (24). However, most research have emphasized provider’ perspective, neglecting how patients independently navigate care-seeking for RTIs. These studies typically rely on secondary data including medical records, pharmacist logs and prescribing audits, offering limited insights into patients’ preferences for seeking antibiotics. For instance, provider data showed over-prescription rates of 29% in China and up to 50% in India and Kenya (25) for presumptive TB and asthma. In Kenyan outpatient clinics, 70.4% of children with upper respiratory tract infections (URTI) received antibiotics at Chogoria and Chuka hospitals (26).

Such datasets provide useful context on prescriber behavior but overlook real-world antibiotic use, especially in community settings where patient decision-making occurs. They also fail to capture how antibiotics are used post-acquisition. Moreover, misinformation, economic hardship, and poor adherence often lead to misuse with only one-third of patients reportedly completing prescribed antibiotics (27). Understanding these antibiotic-seeking or “patient pathways,” as highlighted by (28), is therefore crucial to gaining more comprehensive view of the care-seeking journey, insight that is essential for effectively addressing inappropriate antibiotic use in this population. This gap underscores the need for research that reflects patient’ lived experiences, especially in LMIC setting like Nairobi County, Kenya where antibiotic use for productive coughs remains underexplored despite their potential role in driving antibiotic resistance (29, 30).

Furthermore, sociodemographic factors also shape how patients use antibiotics. According to (31, 32), variables like age, gender, education, and income not only influence adherence to prescribed antibiotics but also shape attitudes and behaviors regarding the acceptance or rejection of antimicrobial interventions (32). Regrettably, their relationship to ABR in patients with productive coughs is poorly understood in Nairobi County. Misuse driven by sociodemographic disparities such as taking antibiotics for viral infections or failing to complete treatment directly contributes to resistance, increasing morbidity, health costs, and mortality.

Therefore, a community-centered approach is vital, aligning with WHO’s framework for AMR control (33), where local-level interventions form the foundation for effective national and global strategies. In response to these knowledge gaps, this article explores how patients with productive coughs in Nairobi County seek, acquire, and use antibiotics. By centering the lived experiences of patients and incorporating sociodemographic context, the research aim to provide actionable insights to inform antimicrobial stewardship (AMS) and reduce the risk of ABR among patients with productive coughs.

## MATERIALS AND METHODS

### Study design and target population

The data reported in this manuscript were abstracted from a larger study entitled “Antibiotic Resistance profiles and molecular basis of Resistance among selected Non-fastidious bacteria recovered from patients with productive coughs in attending selected clinics in Nairobi County, Kenya” which received ethical clearance from the Kenyatta National Hospital–University of Nairobi Ethics and Research Committee (Ref No. KNH-ERC/A/43). The questionnaire data analyzed here focused specifically on antibiotic-seeking behaviors among participants.

This was a facility-based cross-sectional study to assess the patients antibiotic seeking pathways and patterns of antibiotic use among consenting adult patients (>18 years) with productive coughs and attending chest and tuberculosis clinics in Nairobi Kenya. The participants included outpatients presenting at the clinics seeking care for chest conditions.

### Study site

The study was conducted in Nairobi County, Kenya. Nairobi is one of Kenya’s most densely populated urban areas that is marked by a high burden of respiratory diseases driven by contributing factors such as environmental pollution, overcrowding, poor hygiene and sanitation and widespread poverty. These conditions not only increase the risk of respiratory infections but also shape antibiotic seeking behaviors. The study was conducted in four conveniently selected chest and tuberculosis (TB) referral clinics within the County, namely: Mbagathi County Referral Hospital, Rhodes Chest Clinic, Riruta Health Centre, and Kibera D.O. Health Centre. These clinics are specialized centers for diagnosing and treating respiratory conditions such as TB and non-TB pneumonia conditions.

### Eligibility Criteria

#### Inclusion criteria

Any patient aged 18 years or older who presented with a productive cough at the selected chest and tuberculosis clinics and voluntarily consented to participate in the study.

#### Exclusion Criteria

Participants who did not meet the inclusion criteria and could not voluntarily consent to participate in the study.

### Ethical considerations, consenting and confidentiality

Ethical clearance for the study was obtained from the Kenyatta National Hospital-University of Nairobi Ethics Research Committee (KNH-UoN ERC) (Ref No. KNH-ERC/A/43). Permission and approval to conduct the research was also granted by Nairobi County at the selected chest and tuberculosis referral clinics where data collection took place. Participants were prospectively recruited between 30 January 2020 and 29 January 2021. Eligible individuals were approached and invited to participate voluntarily. Those who agreed provided informed consent prior to completing the questionnaire. Participants provided both verbal and written informed consent prior to participation. Only individuals who provided consent were included in the study. To ensure confidentiality, each participant was assigned a unique identification number, which was used on both the consent form and the questionnaire. No personally identifiable information was included in the dataset used for analysis.

### Sampling procedure

#### Data collection

Face-to-face interviews using a structured questionnaire was used to gathered information regarding the social demographic characteristics such as gender, age, marital status, education level, monthly income, and occupation of the participant. The questionnaire also explored antibiotic seeking pathways e.g. through prescription, over-the-counter, self-medication, sharing or using leftover antibiotics or a combination of these pathways. In addition, we collected information on the patterns of antibiotic use with special focus on the overall proportion of participants using antibiotics, diagnosis-based approaches, patterns related to different antibiotic classes and families of antibiotics, frequency of antibiotic usage, completion of antibiotic doses, and use of left over antibiotics as well of sharing of antibiotics.

#### Data analysis

Data was analyzed using R statistical software (version 4.5.1). Univariate analysis was conducted to examine both continuous variables and categorical variables. Frequencies and percentage distributions were used to describe both types of variables, as well as antibiotic-seeking pathways and co-morbidities of the study participants. For bivariate analysis, Pearson’s Chi-square test or Fisher’s exact test was applied to assess the association between dependent variables (e.g., healthcare-seeking pathways) and independent variables (e.g., socio-demographic factors). The odds ratio (OR) and 95% confidence intervals (CI) were calculated to evaluate the strength of these associations. In the multivariate analysis, all independent variables that showed a significant association with the dependent variables in the bivariate analysis were included. Binary logistic regression was used to perform the multivariate analysis, with adjusted odds ratios (AOR) and 95% confidence intervals computed to estimate the strength of the associations.

The threshold for statistical significance was set at p<0.05. Results were presented in narrative, tabular formats.

## RESULTS

### Sociodemographic characteristics of the study participants

Most of the participants were males 65.0% (n=260) and employed 67.0% (n=268). Of those employed, 68.3% (n=183) earned below Ksh 10, 000 (approximately USD 80) per month. More than a third of the participants 35.8% (n=143) had primary/basic education and 37.3% (n=149) had history of smoking, 32.0% (n=128) had history of tuberculosis and 29.8% (n=119) had comorbidities. Among those with comorbidities 51.3% (n=61) had high blood pressure as the most prevalent as shown in table 1 below.

**Table 1:** Descriptive summary of sociodemographic and clinical characteristics of study participants (N=400)

| Variables | n (%) |
| --- | --- |
| **Gender** |  |
| Male | 260 (65.0) |
| Female | 140 (35.0) |
| **Age (years)** |  |
| ≤30 | 100 (25.0) |
| 31-40 | 109 (27.3) |
| 41-50 | 90 (22.5) |
| >50 | 101 (25.3) |
| **Education** |  |
| No formal education | 57 (14.3) |
| Primary/basic education | 143 (35.8) |
| Secondary education | 133 (33.3) |
| Tertiary education | 67 (16.8) |
| **Employment status** |  |
| Employed | 268 (67.0) |
| Unemployed | 132 (33.0) |
| **Gross individual monthly income (among those employed, n=268)** |  |
| <Ksh 10,000 (approx. USD 80) | 183 (68.3) |
| Ksh 10,000 (approx. USD 80) and above | 85 (31.7) |
| **Clinical history** |  |
| History of Tuberculosis (TB) | 128 (32.0) |
| History of smoking | 149 (37.3) |
| Positive Human-Immunodeficiency Virus (HIV) status | 55 (13.8) |
| **Comorbidities** | 119 (29.8) |
| High blood pressure | 61 (51.3) |
| Diabetes | 16 (13.5) |
| Chronic kidney disease | 2(1.7) |

### Antibiotic seeking pathways and patterns of antibiotic use

The majority of participants (86.7%, n=347) reported using antibiotics. Of these, 46.4% (n=161) obtained antibiotics through prescriptions from GP, 31.4% (n=109) acquired them through a combination of GP and OTC pathways, 15.3% (n=53) obtained them solely OTC and 6.9% (n=24) through self-medication pathway. Regarding patterns of usage, 71.1% (n=247) used antibiotics empirically without knowing the cause of their cough, while 28.7% (n=100) had their sputum tested before antibiotic use, with 61% (n=100) of those cases being diagnosed as TB. In terms of frequency, 67.2% (n=233) used antibiotics more than once. The most commonly used antibiotic class was beta-lactams 66.0% (n=229), particularly amoxicillin 90.8% (n=208). Among the 50 participants with leftover antibiotics, all (100%) used them at least once or multiple times. Additionally, 58.3% (n=14) of the participants used antibiotics at least once or multiple times by borrowing or sharing with others as shown in table 2 below.

**Table 2:** Overview of antibiotic use: acquisition routes, usage frequency and patterns, and commonly used antibiotics among study participants with productive coughs (N=400)

| **Variables** | **n (%)** |
| --- | --- |
| **Antibiotic use (overall)** |  |
| Participants reporting antibiotic use | **347 (86.7)** |
| **Antibiotic seeking pathways (n=347)** |  |
| General practitioner (GP) prescription | 161 (46.4) |
| GP and OTC prescription | 109 (31.4) |
| Over the counter (OTC) without prescription | 53 (15.3) |
| Self-medication (borrowing from others) | 24 (6.9) |
| **Patterns of antibiotic use (n=347)** |  |
| Empirical treatment (Sputum analysis not done) | 247 (71.1) |
| Sputum analysis before antibiotic use | 100 (28.7) |
| **Cause of the cough after sputum analysis (n=100)** |  |
| Positive Tuberculosis diagnosis | 61 (61.0) |
| Negative results | 39 (39.0) |
| **Frequency (No. of times/counts) of antibiotic use not in doses** |  |
| Once | 114 (32.9) |
| More than once | 233 (67.2) |
| **Classes of chemotherapeutic regimens used** |  |
| Beta-lactams | 229 (66.0) |
| Anti Tuberculosis | 54 (15.6) |
| Combination of antibiotics and other drugs | 51 (14.7) |
| Macrolides | 8 (2.3) |
| Quinolones | 3 (0.9) |
| Sulfonamides | 2 (0.6) |
| **Most commonly use beta-lactams (n=229)** |  |
| Amoxicillin | 208 (90.8) |
| Amoxicillin/Clavulanic acid | 14 (6.1) |
| Other beta-lactams besides Amoxicillin and Amoxicillin/Clavulanic acid | 7 (3.1) |
| **Most commonly used macrolides (n=8)** |  |
| Azithromycin | 7 (87.5) |
| Clarithromycin | 1 (12.5) |
| **Most commonly used quinolones (n=3)** |  |
| Levofloxacin | 2 (66.7) |
| Ofloxacin | 1 (33.3) |
| **Most commonly sulfa drugs used )n=2)** |  |
| Co-trimoxazole | 2 (100.0) |
| **Availability and use of left-over antibiotics (n=50)** |  |
| Availability of leftover antibiotics within the households | 50 (14.4) |
| Use of leftover Antibiotics | 50 (100.0) |
| **Number of times leftover antibiotics were used (n=50)** |  |
| Once | 24 (48.0) |
| 2-5 times | 19 (38.0) |
| >5 times | 7 (14.0) |
| **Frequency of using shared antibiotics (n=24)** | 24 (100.0) |
| Once | 10 (41.7) |
| 2-5 times | 3 (12.5) |
| >5 times | 1 (4.2) |
| **Not applicable (For participants who borrowed but did not use)** | **10 (41.7)** |
| *Abbreviations: GP- General Practitioner; OTC-Over-the Counter; TB-Tuberculosis* |  |

### Sociodemographic and clinical determinants of antibiotic-seeking pathways

Antibiotic-seeking pathways varied significantly by gender, history of tuberculosis, and income level. Compared to males, females were more likely to engage in self-medication (13.3% vs. 3.2%) and less likely to rely solely on GP prescriptions (41.4% vs. 49.3%; p < 0.01). The history of tuberculosis was associated with greater reliance on GP prescriptions (61.7% vs. 37.4%) and lower engagement in self-medication (3.1% vs. 9.1%; p < 0.01). Among employed users, those earning below Ksh 10,000 (approx. USD 80) were more likely to obtain antibiotics through GP-only routes (52.9% vs. 35.1%), while higher-income earners showed greater use of combined GP and OTC sources (42.9% vs. 24.2%; p = 0.02†). No significant associations were observed with age, education, employment status, smoking, HIV status, comorbidities, or other chronic conditions (Table 3).

**Table 3.** Sociodemographic and clinical associations with antibiotic-seeking pathways among study participants with productive coughs (N=347)

| Variable and categories | GP Rx<br>*N* = 161 | GP+OTC<br>*N* = 109 | OTC only<br>*N* = 53 | Self-medication<br>*N* = 24 | $\chi^2$ | p-value |
| --- | --- | --- | --- | --- | --- | --- |
| **Gender** |  |  |  |  |  |  |
| Male | 108/219 (49.3%) | 67/219 (30.6%) | 37/219 (16.9%) | 7/219 (3.2%) | 14.12 | <0.01 |
| Female | 53/128 (41.4%) | 42/128 (32.8%) | 16/128 (12.5%) | 17/128 (13.3%) |  |  |
| **Age (years)** |  |  |  |  |  |  |
| 30 and below | 41/87 (47.1%) | 26/87 (29.9%) | 13/87 (14.9%) | 7/87 (8.0%) | 4.78 | 0.85 |
| 31-40 | 48/97 (49.5%) | 30/97 (30.9%) | 12/97 (12.4%) | 7/97 (7.2%) |  |  |
| 41-50 | 40/79 (50.6%) | 23/79 (29.1%) | 11/79 (13.9%) | 5/79 (6.3%) |  |  |
| >50 | 32/84 (38.1%) | 30/84 (35.7%) | 17/84 (20.2%) | 5/84 (6.0%) |  |  |
| **Level of education** |  |  |  |  |  |  |
| No formal education | 26/47 (55.3%) | 11/47 (23.4%) | 7/47 (14.9%) | 3/47 (6.4%) | – | 0.91<sup>†</sup> |
| Primary/basic education | 61/130 (46.9%) | 43/130 (33.1%) | 19/130 (14.6%) | 7/130 (5.4%) |  |  |
| Secondary education | 49/110 (44.5%) | 35/110 (31.8%) | 16/110 (14.5%) | 10/110 (9.1%) |  |  |
| College/university | 25/60 (41.7%) | 20/60 (33.3%) | 11/60 (18.3%) | 4/60 (6.7%) |  |  |
| **Employment status** |  |  |  |  |  |  |
| Employed | 110/234 (47.0%) | 71/234 (30.3%) | 38/234 (16.2%) | 15/234 (6.4%) | 1.02 | 0.80 |
| Unemployed | 51/113 (45.1%) | 38/113 (33.6%) | 15/113 (13.3%) | 9/113 (8.0%) |  |  |
| **Gross individual monthly income (among employed users, n=234)** |  |  |  |  |  |  |
| Below Ksh 10,000 (approx. USD 80) | 83/157 (52.9%) | 38/157 (24.2%) | 25/157 (15.9%) | 11/157 (7.0%) | – | 0.02† |
| Ksh 10,000 (approx. USD 80) and above | 27/77 (35.1%) | 33/77 (42.9%) | 13/77 (16.9%) | 4/77 (5.2%) |  |  |
| **History of TB** |  |  |  |  |  |  |
| Yes | 79/128 (61.7%) | 31/128 (24.2%) | 14/128 (10.9%) | 4/128 (3.1%) | 20.31 | <0.01 |
| No | 82/219 (37.4%) | 78/219 (35.6%) | 39/219 (17.8%) | 20/219 (9.1%) |  |  |
| **Had history of smoking** |  |  |  |  |  |  |
| No | 104/225 (46.2%) | 69/225 (30.7%) | 34/225 (15.1%) | 18/225 (8.0%) | 1.21 | 0.75 |
| Yes | 57/122 (46.7%) | 40/122 (32.8%) | 19/122 (15.6%) | 6/122 (4.9%) |  |  |
| **HIV status** |  |  |  |  |  |  |
| Negative | 134/294 (45.6%) | 93/294 (31.6%) | 46/294 (15.6%) | 21/294 (7.1%) |  |  |
| Positive | 27/53 (50.9%) | 16/53 (30.2%) | 7/53 (13.2%) | 3/53 (5.7%) | – | 0.93† |
| No response |  |  |  |  |  |  |
| **Comorbidity** |  |  |  |  |  |  |
| No | 110/239 (46.0%) | 75/239 (31.4%) | 41/239 (17.2%) | 13/239 (5.4%) | 4.22 | 0.24 |
| Yes | 51/108 (47.2%) | 34/108 (31.5%) | 12/108 (11.1%) | 11/108 (10.2%) |  |  |
| **Type of comorbidity** |  |  |  |  |  |  |
| **Had high blood pressure** |  |  |  |  |  |  |
| No | 138/293 (47.1%) | 92/293 (31.4%) | 47/293 (16.0%) | 16/293 (5.5%) | – | 0.12† |
| Yes | 23/54 (42.6%) | 17/54 (31.5%) | 6/54 (11.1%) | 8/54 (14.8%) |  |  |
| **Had diabetes** |  |  |  |  |  |  |
| No | 153/334 (45.8%) | 105/334 (31.4%) | 53/334 (15.9%) | 23/334 (6.9%) | – | 0.39† |
| Yes | 8/13 (61.5%) | 4/13 (30.8%) | 0/13 (0.0%) | 1/13 (7.7%) |  |  |
| **Note:** |  |  |  |  |  |  |
| Cells show n/N (%) within each row category. |  |  |  |  |  |  |
| P-values are from Pearson's $\chi^2$; when any expected count < 5, Fisher's exact test (Monte Carlo as needed) is used and marked with † ($\chi^2$ omitted). | | | | | | |
| Percentages rounded to 1 dp; p-values to 2 dp (half-up rounding). |  |  |  |  |  |  |
| Analysis restricted to antibiotic users only; income shown among employed users only. |  |  |  |  |  |  |
| Abbreviations: GP Rx = General Practitioners (GP) prescription; GP+OTC = GP and OTC prescription; OTC only = Over the counter only; Self-medication = Self-medication by borrowing from others. |  |  |  |  |  |  |

In the crude multinomial model (reference category: GP prescription only), several factors were significantly associated with alternative antibiotic-seeking pathways. Female participants had significantly higher relative risk of self-medicating compared to males (RRR: 4.95; 95% CI: 1.93–12.66; p < 0.01), but did not differ significantly in their use of GP plus OTC or OTC-only sources. Participants with a history of tuberculosis were significantly less likely to use any alternative pathway compared to GP-only: GP plus OTC (RRR: 0.41; 95% CI: 0.25–0.69; p < 0.01); OTC only (RRR: 0.37; 95% CI: 0.19–0.74; p < 0.01); Self-medication (RRR: 0.21; 95% CI: 0.07–0.63; p < 0.01). Among employed users, higher income (≥ Ksh 10,000 (approx. USD 80) was associated with greater likelihood of using both GP and OTC sources (RRR: 2.67; 95% CI: 1.41–5.05; p < 0.01) compared to lower-income users. Participants with high blood pressure had increased relative risk of self-medication compared to those without (RRR: 3.00; 95% CI: 1.15–7.81; p = 0.02). No statistically significant associations were observed for age, education, employment status, smoking, HIV status, comorbidity, or diabetes across any pathway (Table 4).

**Table 4.** Crude multinomial logistic regression analysis of sociodemographic and clinical factors associated with antibiotic-seeking pathways among study participants with productive coughs (n=347)

| Variable and categories | GP+OTC RRR (95% CI) | p-value | OTC only RRR (95% CI) | p-value | Self-medication RRR (95% CI) | p-value |
| --- | --- | --- | --- | --- | --- | --- |
| **Gender** |  |  |  |  |  |  |
| Male | Ref |  | Ref |  | Ref |  |
| Female | 1.28 (0.77–2.12) | 0.34 | 0.88 (0.45–1.73) | 0.71 | 4.95 (1.93–12.66) | <0.01** |
| **Age (years)** |  |  |  |  |  |  |
| 30 and below | Ref |  | Ref |  | Ref |  |
| 31–40 | 0.99 (0.50–1.93) | 0.97 | 0.79 (0.32–1.92) | 0.60 | 0.85 (0.28–2.64) | 0.78 |
| 41–50 | 0.91 (0.45–1.84) | 0.79 | 0.87 (0.35–2.16) | 0.76 | 0.73 (0.21–2.50) | 0.62 |
| >50 | 1.48 (0.73–2.98) | 0.27 | 1.68 (0.71–3.95) | 0.24 | 0.92 (0.27–3.15) | 0.89 |
| **Level of education** |  |  |  |  |  |  |
| No formal education | Ref |  | Ref |  | Ref |  |
| Primary/basic education | 1.67 (0.74–3.73) | 0.21 | 1.16 (0.43–3.08) | 0.77 | 0.99 (0.24–4.15) | 0.99 |
| Secondary education | 1.69 (0.74–3.86) | 0.21 | 1.21 (0.44–3.32) | 0.71 | 1.77 (0.45–7.00) | 0.42 |
| College/university | 1.89 (0.76–4.74) | 0.17 | 1.63 (0.55–4.89) | 0.38 | 1.39 (0.28–6.83) | 0.69 |
| **Employment status** |  |  |  |  |  |  |
| Unemployed | Ref |  | Ref |  | Ref |  |
| Employed | 0.87 (0.52–1.45) | 0.59 | 1.17 (0.59–2.33) | 0.64 | 0.77 (0.32–1.88) | 0.57 |
| **Gross individual monthly income (among employed users, n=234)** |  |  |  |  |  |  |
| Below KSH 10,000 (approx. USD 80) | Ref |  | Ref |  | Ref |  |
| Ksh 10,000 (approx. USD 80) and above | 2.67 (1.41–5.05) | <0.01** | 1.60 (0.72–3.55) | 0.25 | 1.12 (0.33–3.80) | 0.86 |
| **Had history of TB** |  |  |  |  |  |  |
| No | Ref |  | Ref |  | Ref |  |
| Yes | 0.41 (0.25–0.69) | <0.01** | 0.37 (0.19–0.74) | <0.01** | 0.21 (0.07–0.63) | <0.01** |
| **Had history of smoking** |  |  |  |  |  |  |
| No | Ref |  | Ref |  | Ref |  |
| Yes | 1.06 (0.64–1.75) | 0.83 | 1.02 (0.53–1.95) | 0.95 | 0.61 (0.23–1.62) | 0.32 |
| **Positive HIV status** |  |  |  |  |  |  |
| Negative | Ref |  | Ref |  | Ref |  |
| Positive | 0.85 (0.44–1.67) | 0.65 | 0.76 (0.31–1.85) | 0.54 | 0.71 (0.20–2.55) | 0.60 |
| **Comorbidity** |  |  |  |  |  |  |
| No | Ref |  | Ref |  | Ref |  |
| Yes | 0.98 (0.58–1.65) | 0.93 | 0.63 (0.31–1.30) | 0.21 | 1.82 (0.77–4.35) | **0.17** |
| **Had high blood pressure** |  |  |  |  |  |  |
| No | Ref |  | Ref |  | Ref |  |
| Yes | 1.11 (0.56–2.19) | 0.77 | 0.77 (0.29–2.00) | 0.59 | 3.00 (1.15–7.81) | **0.02\*\*** |
| **Had diabetes** |  |  |  |  |  |  |
| No | Ref |  | Ref |  | Ref |  |
| Yes | 0.73 (0.21–2.49) | 0.61 | 0.00 (0.00–177550713.73) | 0.63 | 0.83 (0.10–6.96) | 0.86 |

**Note:**
*Dependent variable: Antibiotic-seeking pathways (multinomial; baseline = GP Rx); users only.*
*RRR = Relative Risk Ratio from univariable multinomial logistic regression; Wald P-values.*
*Bold P-values indicate $P < 0.20$ retained for adjusted model; \*\* marks 95% significance.*
*Income among employed users only.*
*Abbreviations: GP Rx = General Practitioners prescription; GP+OTC = GP and OTC prescription; OTC only = Over the counter only;*
*Self-medication = Self-medication by borrowing from others.*

In the adjusted model (reference category: GP prescription only), two variables remained significantly associated with alternative antibiotic-seeking pathways: gender and history of tuberculosis. Female participants had significantly higher relative risk of self-medication compared to males (aRRR: 3.83; 95% CI: 1.46–10.03; p < 0.01), but did not differ significantly in their likelihood of using GP+OTC or OTC-only sources. Participants with a history of TB were significantly less likely to use any alternative antibiotic-seeking pathway: GP+OTC (aRRR: 0.42; 95% CI: 0.25–0.71; p < 0.01); OTC only (aRRR: 0.35; 95% CI: 0.17–0.70; p < 0.01); Self-medication (aRRR: 0.28; 95% CI: 0.09–0.89; p = 0.03). No other variables met the threshold for inclusion in the final model after backward elimination (Table 5).

**Table 5.**
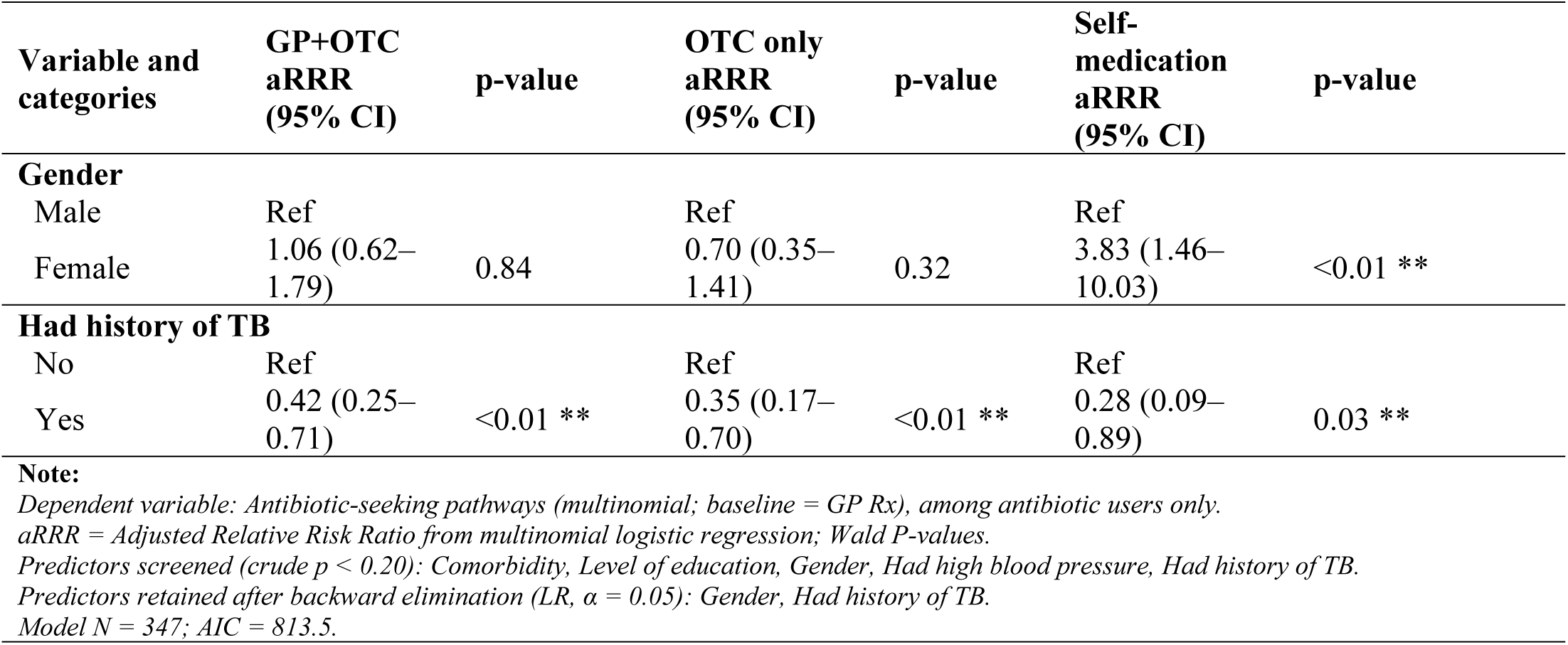
Adjusted multinomial associations between gender, and TB history, and antibiotic-seeking pathways among study participants with productive coughs (n = 347)

| Variable and categories | GP+OTC<br>aRRR<br>(95% CI) | p-value | OTC only<br>aRRR<br>(95% CI) | p-value | Self-medication<br>aRRR<br>(95% CI) | p-value |
| --- | --- | --- | --- | --- | --- | --- |
| **Gender** |  |  |  |  |  |  |
| Male | Ref |  | Ref |  | Ref |  |
| Female | 1.06 (0.62–1.79) | 0.84 | 0.70 (0.35–1.41) | 0.32 | 3.83 (1.46–10.03) | <0.01 ** |
| **Had history of TB** |  |  |  |  |  |  |
| No | Ref |  | Ref |  | Ref |  |
| Yes | 0.42 (0.25–0.71) | <0.01 ** | 0.35 (0.17–0.70) | <0.01 ** | 0.28 (0.09–0.89) | 0.03 ** |

**Note:** Dependent variable: Antibiotic-seeking pathways (multinomial; baseline = GP Rx), among antibiotic users only. aRRR = Adjusted Relative Risk Ratio from multinomial logistic regression; Wald P-values. Predictors screened (crude $p < 0.20$ ): Comorbidity, Level of education, Gender, Had high blood pressure, Had history of TB. Predictors retained after backward elimination (LR, $\alpha = 0.05$ ): Gender, Had history of TB. Model $N = 347$ ; AIC = 813.5.

**Table 6.**
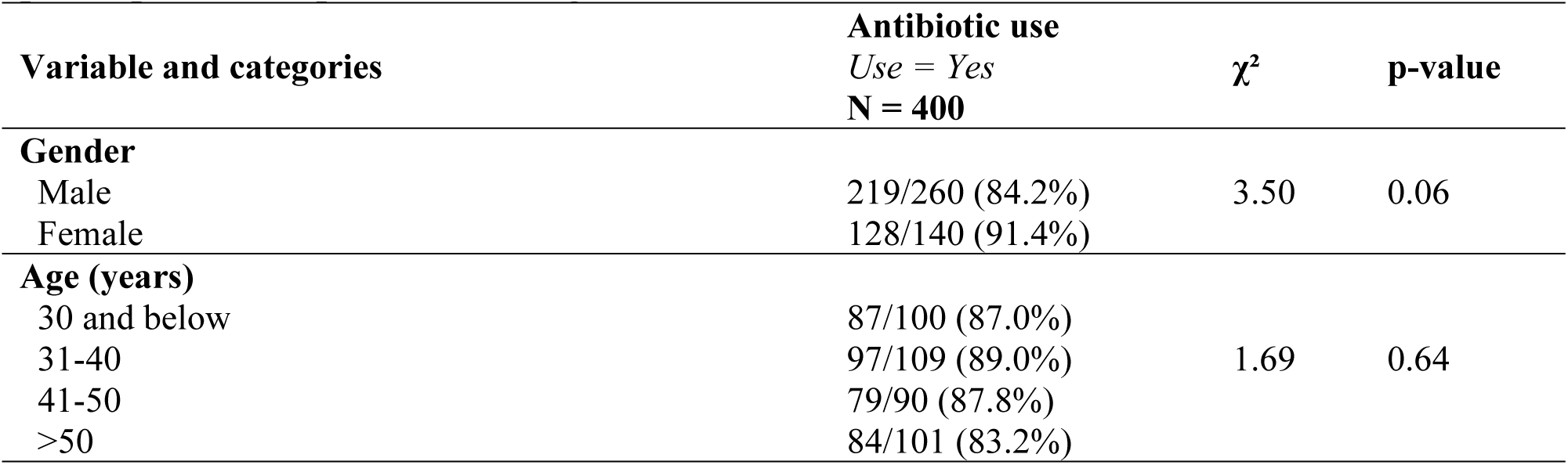
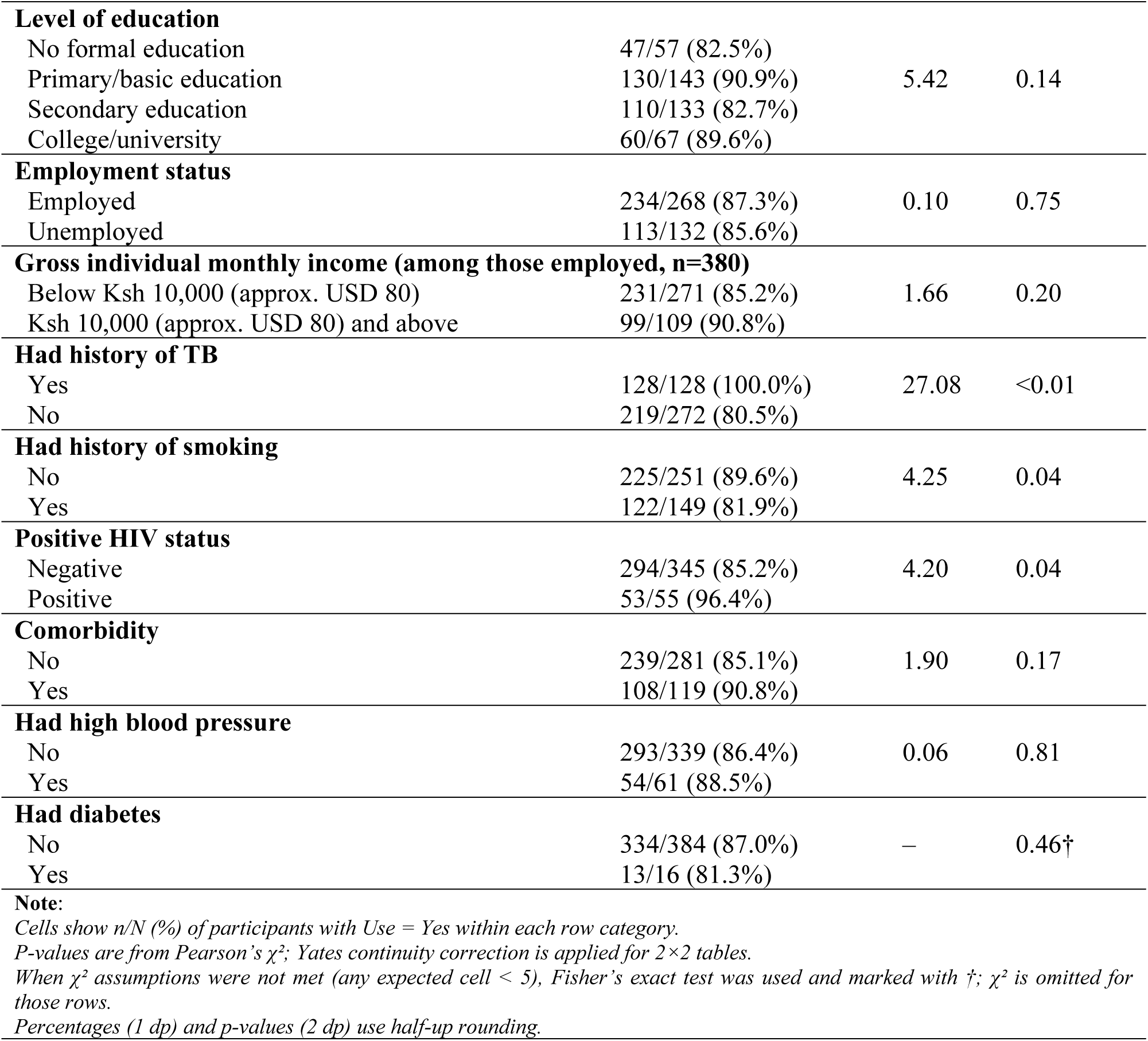
Sociodemographic and clinical determinants of antibiotic use among study participants with productive coughs (n = 400)

| Variable and categories | Antibiotic use<br>Use = Yes<br>N = 400 | $\chi^2$ | p-value |
| --- | --- | --- | --- |
| **Gender** |  |  |  |
| Male | 219/260 (84.2%) | 3.50 | 0.06 |
| Female | 128/140 (91.4%) |  |  |
| **Age (years)** |  |  |  |
| 30 and below | 87/100 (87.0%) | 1.69 | 0.64 |
| 31-40 | 97/109 (89.0%) |  |  |
| 41-50 | 79/90 (87.8%) |  |  |
| >50 | 84/101 (83.2%) |  |  |
| **Level of education** |  |  |  |
| No formal education | 47/57 (82.5%) |  |  |
| Primary/basic education | 130/143 (90.9%) | 5.42 | 0.14 |
| Secondary education | 110/133 (82.7%) |  |  |
| College/university | 60/67 (89.6%) |  |  |
| **Employment status** |  |  |  |
| Employed | 234/268 (87.3%) | 0.10 | 0.75 |
| Unemployed | 113/132 (85.6%) |  |  |
| **Gross individual monthly income (among those employed, n=380)** |  |  |  |
| Below Ksh 10,000 (approx. USD 80) | 231/271 (85.2%) | 1.66 | 0.20 |
| Ksh 10,000 (approx. USD 80) and above | 99/109 (90.8%) |  |  |
| **Had history of TB** |  |  |  |
| Yes | 128/128 (100.0%) | 27.08 | <0.01 |
| No | 219/272 (80.5%) |  |  |
| **Had history of smoking** |  |  |  |
| No | 225/251 (89.6%) | 4.25 | 0.04 |
| Yes | 122/149 (81.9%) |  |  |
| **Positive HIV status** |  |  |  |
| Negative | 294/345 (85.2%) | 4.20 | 0.04 |
| Positive | 53/55 (96.4%) |  |  |
| **Comorbidity** |  |  |  |
| No | 239/281 (85.1%) | 1.90 | 0.17 |
| Yes | 108/119 (90.8%) |  |  |
| **Had high blood pressure** |  |  |  |
| No | 293/339 (86.4%) | 0.06 | 0.81 |
| Yes | 54/61 (88.5%) |  |  |
| **Had diabetes** |  |  |  |
| No | 334/384 (87.0%) | – | 0.46† |
| Yes | 13/16 (81.3%) |  |  |

**Note:**
Cells show n/N (%) of participants with Use = Yes within each row category.
P-values are from Pearson's $\chi^2$ ; Yates continuity correction is applied for 2×2 tables.
When $\chi^2$ assumptions were not met (any expected cell < 5), Fisher's exact test was used and marked with †; $\chi^2$ is omitted for those rows.
Percentages (1 dp) and p-values (2 dp) use half-up rounding.

### Sociodemographic and clinical determinants of antibiotic use

Among the 400 participants, antibiotic use was high overall (87.3%) but varied across clinical and sociodemographic factors. Antibiotic use was significantly higher among participants with a history of tuberculosis (100.0% vs. 80.5%, p < 0.01), those with a positive HIV status (96.4% vs. 85.2%, p = 0.04), and non-smokers compared to smokers (89.6% vs. 81.9%, p = 0.04). No statistically significant associations were observed between antibiotic use and gender (p = 0.06), age group (p = 0.64), level of education (p = 0.14), employment status (p = 0.75), income level (p = 0.20), comorbidity presence (p = 0.17), hypertension status (p = 0.81), or diabetes status (p = 0.46) (Tale 6).

A crude logistic regression model was fitted to further explore associations between predictor variables and antibiotic use. Female participants had twice the odds of antibiotic use compared to males (cOR: 2.00; 95% CI: 1.04–4.10; p = 0.05). Participants with a history of TB had significantly higher odds of antibiotic use (cOR: 185.99; 95% CI: 1.51–22881.78; p = 0.03), as did those with positive HIV status (cOR: 4.60; 95% CI: 1.37–28.63; p = 0.04). Conversely, a history of smoking was associated with lower odds of antibiotic use (cOR: 0.52; 95% CI: 0.29–0.94; p = 0.03). Other variables, including age, education level, employment, income, comorbidity status, hypertension, and diabetes, showed no statistically significant associations at the 95% confidence level, though variables with p < 0.20 were retained for adjusted analysis (Table 7).

**Table 7.** Crude associations between sociodemographic and clinical factors and antibiotic use among study participants with productive coughs (n = 400)

| Variable and categories | cOR (95% CI) | p-value |
| --- | --- | --- |
| **Gender** |  |  |
| Male | Ref |  |
| Female | 2.00 (1.04–4.10) | **0.05 \*\*** |
| **Age (years)** |  |  |
| 30 and below | Ref |  |
| 31-40 | 1.21 (0.52–2.82) | 0.66 |
| 41-50 | 1.07 (0.45–2.58) | 0.87 |
| >50 | 0.74 (0.33–1.61) | 0.45 |
| **Level of education** |  |  |
| No formal education | Ref |  |
| Primary/basic education | 2.13 (0.86–5.17) | **0.10** |
| Secondary education | 1.02 (0.43–2.25) | 0.97 |
| College/university | 1.82 (0.65–5.37) | 0.26 |
| **Employment status** |  |  |
| Unemployed | Ref |  |
| Employed | 1.16 (0.62–2.10) | 0.64 |
| **Gross individual monthly income (among those employed, n=268)** |  |  |
| Below Ksh 10,000 (approx. USD 80) | Ref |  |
| Ksh 10,000 (approx. USD 80) and above | 1.59 (0.72–3.92) | 0.28 |
| **Had history of TB** |  |  |
| No | Ref |  |
| Yes | 185.99 (1.51–22881.78) | **0.03 \*\*** |
| **Had history of smoking** |  |  |
| No | Ref |  |
| Yes | 0.52 (0.29–0.94) | **0.03 \*\*** |
| **Positive HIV status** |  |  |
| Negative | Ref |  |
| Positive | 4.60 (1.37–28.63) | **0.04 \*\*** |
| **Comorbidity** |  |  |
| No | Ref |  |
| Yes | 1.73 (0.88–3.64) | **0.13** |
| **Had high blood pressure** |  |  |
| No | Ref |  |
| Yes | 1.21 (0.55–3.06) | 0.66 |
| **Had diabetes** |  |  |
| No | Ref |  |
| Yes | 0.65 (0.20–2.90) | 0.51 |
| **Note:** |  |  |
| *Dependent variable: Antibiotic use (Use = Yes vs No).* |  |  |
| *cOR = Crude Odds Ratio from univariable logistic regression;* |  |  |
| *P-values are from Wald tests.* |  |  |
| *Bold p-values indicate predictors with $p < 0.20$, retained for adjusted analysis; indicates significance at the 95% confidence level. Models with separation were refitted using bias-reduced logistic regression (brglm2, AS_median).* |  |  |
| *Ref indicates the reference category in the cOR column.* |  |  |

In the adjusted logistic regression model, two clinical factors remained significantly associated with antibiotic use. Participants with a history of TB had substantially higher odds of antibiotic use (aOR: 258.64; 95% CI: 2.13–31,469.18; p = 0.02), while those with a history of smoking had significantly lower odds (aOR: 0.32; 95% CI: 0.17–0.59; p < 0.01). No sociodemographic variables were retained in the final model after backward elimination (Table 8).

**Table 8.**
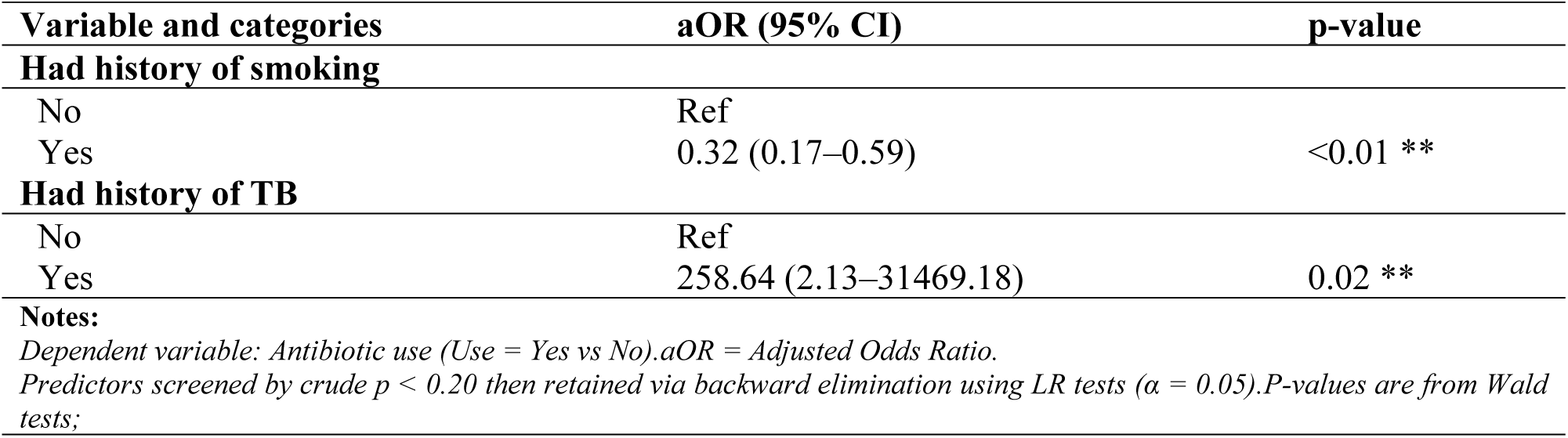
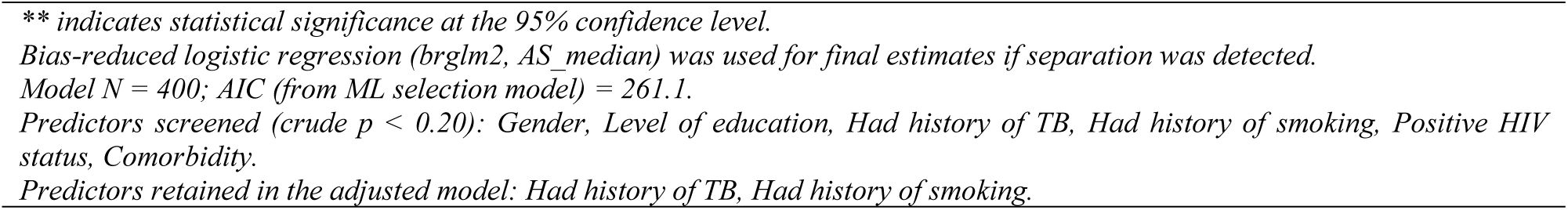
Adjusted associations between clinical factors and antibiotic use among study participants with productive coughs (n = 400)

| Variable and categories | aOR (95% CI) | p-value |
| --- | --- | --- |
| **Had history of smoking** |  |  |
| No | Ref |  |
| Yes | 0.32 (0.17–0.59) | <0.01 ** |
| **Had history of TB** |  |  |
| No | Ref |  |
| Yes | 258.64 (2.13–31469.18) | 0.02 ** |
| **Notes:** |  |  |
| *Dependent variable: Antibiotic use (Use = Yes vs No).aOR = Adjusted Odds Ratio.* |  |  |
| *Predictors screened by crude $p < 0.20$ then retained via backward elimination using LR tests ($\alpha = 0.05$).P-values are from Wald tests;* |  |  |

*\*\* indicates statistical significance at the 95% confidence level.*
*Bias-reduced logistic regression (brglm2, AS\_median) was used for final estimates if separation was detected.*
*Model N = 400; AIC (from ML selection model) = 261.1.*
*Predictors screened (crude $p < 0.20$ ): Gender, Level of education, Had history of TB, Had history of smoking, Positive HIV status, Comorbidity.*
*Predictors retained in the adjusted model: Had history of TB, Had history of smoking.*

### Sociodemographic and clinical determinants of frequency of antibiotic use

Among participants who reported antibiotic use, frequency of use differed significantly by several characteristics. Female participants were more likely than males to report using antibiotics more than once (80.5% vs. 59.4%, p < 0.01). Those with a gross monthly income of Ksh 10,000 (approx. USD 80) and above were also more likely to report repeat use compared to those earning less (75.8% vs. 61.9%, p = 0.02). The history of tuberculosis was significantly associated with higher frequency of antibiotic use (57.8% vs. 27.4% reporting single use, p < 0.01), and a similar trend was observed for participants with high blood pressure (79.6% vs. 64.8%, p = 0.05). Other variables including age, education level, employment status, smoking history, HIV status, comorbidity presence, and diabetes were not significantly associated with frequency of antibiotic use at the bivariate level (Table 9).

**Table 9.** Sociodemographic and clinical determinants of frequency of antibiotic use among study participants with productive coughs (N = 347).

| Variable and categories | Frequency of antibiotic use | | $\chi^2$ | p-value |
| --- | --- | --- | --- | --- |
|  | Once<br>N (%) | More than once<br>N (%) |  |  |
| **Gender** |  |  |  |  |
| Male | 89 (40.6%) | 130 (59.4%) | 15.37 | <0.01 |
| Female | 25 (19.5%) | 103 (80.5%) |  |  |
| **Age (years)** |  |  |  |  |
| 30 and below | 28 (32.2%) | 59 (67.8%) | 5.02 | 0.17 |
| 31-40 | 40 (41.2%) | 57 (58.8%) |  |  |
| 41-50 | 24 (30.4%) | 55 (69.6%) |  |  |
| >50 | 22 (26.2%) | 62 (73.8%) |  |  |
| **Level of education** |  |  |  |  |
| No formal education | 13 (27.7%) | 34 (72.3%) | 3.03 | 0.39 |
| Primary/basic education | 48 (36.9%) | 82 (63.1%) |  |  |
| Secondary education | 31 (28.2%) | 79 (71.8%) |  |  |
| College/university | 22 (36.7%) | 38 (63.3%) |  |  |
| **Employment status** |  |  |  |  |
| Employed | 80 (34.2%) | 154 (65.8%) | 0.41 | 0.52 |
| Unemployed | 34 (30.1%) | 79 (69.9%) |  |  |
| **Gross individual monthly income (among employed users, n=330)** |  |  |  |  |
| Below Ksh 10,000 (approx. USD 80) | 88 (38.1%) | 143 (61.9%) | 5.33 | 0.02 |
| Ksh 10,000 (approx. USD 80) and above | 24 (24.2%) | 75 (75.8%) |  |  |
| **Had history of TB** |  |  |  |  |
| Yes | 54 (42.2%) | 74 (57.8%) | 7.35 | <0.01 |
| No | 60 (27.4%) | 159 (72.6%) |  |  |
| **Had history of smoking** |  |  |  |  |
| No | 71 (31.6%) | 154 (68.4%) | 0.34 | 0.56 |
| Yes | 43 (35.2%) | 79 (64.8%) |  |  |
| **Positive HIV status** |  |  |  |  |
| Negative | 96 (32.7%) | 198 (67.3%) | 0.00 | 0.98 |
| Positive | 18 (34.0%) | 35 (66.0%) |  |  |
| **Comorbidity** |  |  |  |  |
| No | 84 (35.1%) | 155 (64.9%) | 1.51 | 0.22 |
| Yes | 30 (27.8%) | 78 (72.2%) |  |  |
| **Had high blood pressure** |  |  |  |  |
| No | 103 (35.2%) | 190 (64.8%) | 3.87 | 0.05 |
| Yes | 11 (20.4%) | 43 (79.6%) |  |  |
| **Had diabetes** |  |  |  |  |
| No | 111 (33.2%) | 223 (66.8%) | — | 0.56† |
| Yes | 3 (23.1%) | 10 (76.9%) |  |  |

Note:
Cells shown (row %) across the two frequency categories within each row.
P-values are from Pearson's $\chi^2$ ; Yates continuity correction is applied for 2×2 tables.
When $\chi^2$ assumptions were not met (any expected cell < 5), Fisher's exact test was used and marked with †; $\chi^2$ is omitted for those rows.
Percentages (1 dp) and p-values (2 dp) use half-up rounding.
Income is evaluated among employed users only.

A crude logistic regression model revealed several factors associated with frequent antibiotic use. Female participants had nearly three times the odds of reporting multiple antibiotic uses compared to males (cOR: 2.82; 95% CI: 1.71–4.79; p < 0.01). Participants earning Ksh 10,000 (approx. USD 80) or more per month had higher odds of frequent use compared to those earning less (cOR: 1.94; 95% CI: 1.07–3.63; p = 0.03). A history of tuberculosis was associated with lower odds of frequent use (cOR: 0.52; 95% CI: 0.33–0.82; p < 0.01), while high blood pressure was linked to increased odds of repeated antibiotic use (cOR: 2.12; 95% CI: 1.08–4.48; p = 0.04). Other variables showed no significant associations at the 95% confidence level, although predictors with p < 0.20 were retained for multivariable analysis (Table 10).

**Table 10.** Crude associations between sociodemographic and clinical factors and frequency of antibiotic use among study participants with productive coughs (n = 347)

| Variable and categories | cOR (95% CI) | p-value |
| --- | --- | --- |
| **Gender** |  |  |
| Male | Ref |  |
| Female | 2.82 (1.71–4.79) | **<0.01 \*\*** |
| **Age (years)** |  |  |
| 30 and below | Ref |  |
| 31-40 | 0.68 (0.37–1.23) | 0.20 |
| 41-50 | 1.09 (0.56–2.11) | 0.80 |
| >50 | 1.34 (0.69–2.61) | 0.39 |
| **Level of education** |  |  |
| No formal education | Ref |  |
| Primary/basic education | 0.65 (0.31–1.33) | 0.25 |
| Secondary education | 0.97 (0.44–2.06) | 0.95 |
| College/university | 0.66 (0.28–1.50) | 0.33 |
| **Employment status** |  |  |
| Unemployed | Ref |  |
| Employed | 0.83 (0.51–1.34) | 0.45 |
| **Gross individual monthly income (among employed users, n=234)** |  |  |
| Below Ksh 10,000 (approx. USD 80) | Ref |  |
| Ksh 10,000 (approx. USD 80) and above | 1.94 (1.07–3.63) | **0.03 \*\*** |
| **Had history of TB** |  |  |
| No | Ref |  |
| Yes | 0.52 (0.33–0.82) | **<0.01 \*\*** |
| **Had history of smoking** |  |  |
| No | Ref |  |
| Yes | 0.85 (0.53–1.35) | 0.48 |
| **Positive HIV status** |  |  |
| Negative | Ref |  |
| Positive | 0.94 (0.51–1.78) | 0.85 |
| **Comorbidity** |  |  |
| No | Ref |  |
| Yes | 1.41 (0.86–2.34) | **0.18** |
| **Had high blood pressure** |  |  |
| No | Ref |  |
| Yes | 2.12 (1.08–4.48) | **0.04 \*\*** |
| **Had diabetes** |  |  |
| No | Ref |  |
| Yes | 1.66 (0.50–7.51) | 0.45 |

**Note:**
Outcome coded as More than once (1) vs once (0); analysis restricted to antibiotic users (excludes 'Never took antibiotics').
cOR = Crude Odds Ratio from univariable logistic regression; P-values are from Wald tests.
Bold p-values indicate predictors with $p < 0.20$ retained for adjusted analysis; \*\* indicates significance at the 95% confidence level.
Models exhibiting separation were refit with bias-reduced logistic regression (brglm2, AS\_median).
Income is evaluated among employed users only.

In the adjusted model, female participants remained significantly more likely to report frequent antibiotic use (aOR: 2.55; 95% CI: 1.53–4.37; p < 0.01). Additionally, a history of tuberculosis was independently associated with reduced odds of frequent use (aOR: 0.62; 95% CI: 0.38–1.00; p = 0.05). Variables such as comorbidity and high blood pressure, while significant at the crude level, were not retained in the final model after backward elimination (Table 11).

**Table 11.** Adjusted associations between clinical factors and frequency of antibiotic use among study participants with productive coughs (N = 347).

| Variable and categories | aOR (95% CI) | p-value |
| --- | --- | --- |
| **Gender** |  |  |
| Male | Ref |  |
| Female | 2.55 (1.53–4.37) | <0.01 ** |
| **Had history of TB** |  |  |
| No | Ref |  |
| Yes | 0.62 (0.38–1.00) | 0.05 ** |
| **Note:** |  |  |
| *Dependent variable: Frequency of antibiotic use (More than once vs once), among antibiotic users only.* |  |  |
| *Predictors screened (crude $p < 0.20$): Gender, Had history of TB, Comorbidity, Had high blood pressure.* |  |  |
| *Predictors retained in the adjusted model: Gender, Had history of TB.* |  |  |
| *P-values are from Wald tests; \*\* indicates statistical significance at the 95% confidence level.* |  |  |
| *Bias-reduced logistic regression (brglm2, AS_median) was used for final estimates if separation was detected.* |  |  |
| *Model N = 347; AIC (from ML selection model) = 424.4.* |  |  |

## DISCUSSION

This study explored the patterns and pathways through which individuals with productive coughs in Nairobi County seek and consume antibiotics. It highlights the roles of formal healthcare antibiotic access, OTC purchases, and self-medication, and examines how sociodemographic and health-related factors influence these behaviors. Drawing on both local and international studies, this addresses the widespread misuse of antibiotics, often driven by misconceptions, poor regulation, and limited diagnostic support, which contributes significantly to the growing challenge of antimicrobial resistance. The findings underscore the urgent need for improved antimicrobial stewardship and better access to accurate diagnostic services in resource-limited settings.

In this study, patients with productive coughs sought antibiotics through various pathways, which differed among study participants. The most common route, chosen by 46.4% of participants (161 individuals), was obtaining antibiotics from General Practitioners (GPs). This findings aligns with (25), who reported that antibiotics prescription by GPs as the most common source of antibiotics, with rates ranging from 29% in China to 50% in India and Kenya, often for conditions like presumptive TB and asthma. Similarly, in South Africa, (34) found that GPs were the main source of antibiotics, prescribing them 55.5% of the time for RTIs in more than 70% of the case. Elsewhere, (35) reported an even higher antibiotic prescription rate of 90% for RTIs in Kenya, with most antibiotics issued through valid prescription from a GP. This suggests that RTIs drives a significant number of primary healthcare visits (36).

Complementary to these findings, studies also show that individuals with respiratory symptoms frequently seek care at formal medical institution. In China, 50.92% of patients with acute respiratory infections sought treatment at medical institutions (37) while in Kenya, 37–43% of patients with mild respiratory illnesses and 18–24% of those with severe pneumonia visited primary health care facilities (38). Similarly, in Ethiopia, 80% of individuals with respiratory illnesses reported visiting health posts, health centers, or hospital (39) while those with chronic cough due to tuberculosis sought diagnosis in health facilities in Angola (40).

Unfortunately, GPs often overprescribe antibiotics for RTIs, many of which are unnecessary and approximately 90.25% of them are reported to be inappropriately prescribed (41). For example, studies have shown that 42.2% of patients with bronchitis, 31.9% with nonspecific acute upper respiratory infections, and 25.9% with pharyngitis were prescribed antibiotics unnecessarily (42, 43). Similarly, over 30%–50% of all outpatients received unnecessary or inappropriate antibiotics for viral upper respiratory infections in the Unites states (44). Frequent inappropriate prescription of antibiotics may contribute to antibiotic resistance, a public health crisis that is especially concerning in Kenya, where the rate of antimicrobial resistance is rising rapidly (45). In fact, broader analysis estimate that 40–80% of these antibiotics are unnecessarily prescribed (46), and up to 50% are misused (47). One reason for this trend is the diagnostic uncertainty where 30–40% of RTIs remain undiagnosed (2) and many are not bacteriologically confirmed before prescribing antibiotics making it difficult for clinicians and GP to determine whether antibiotics are truly needed.

In addition to the prescription pathway, 53 participants (15.3%) sought antibiotics through the OTC route from pharmacies without a prescription. This finding aligns with the recently reported prevalence of OTC antibiotic use in Sub-Saharan Africa, ranging from 8% in Zimbabwe to 94% in Uganda (48). In Nairobi County, the practice of purchasing no-prescribed antibiotics for RTIs at a relatively high rate of (15.3%) mirrors but exceeds the 5.9% reported by (35) for dispensing antibiotics without prescriptions in Kenya.

This discrepancy may be attributed to variations in study design, scope, and data collection methods. For instance, (35) relied on secondary data drawn from 222 pharmacy records, which may have potentially underestimated informal or unrecorded dispensing practices. In contrast, the present study utilized a larger sample of 400 participants presenting with productive coughs, thereby capturing a broader spectrum of antibiotic-seeking behaviors. Additionally, the higher rate observed may reflect ongoing challenges, such as the fact that over 70% of pharmacists in Kenya reportedly do not require prescriptions before dispensing antibiotics (49), as well as the weak enforcement of existing pharmaceutical regulations (45).

This is reflected more broadly in Sub-Saharan Africa, where 69% of patients who visited a pharmacy and either requested antibiotics or consulted pharmacy staff for URTI symptoms received antibiotics without a prescription (48). Additionally, 21.5% of patients with acute respiratory illness and 20.8% of those with severe pneumonia in Kenya reported purchasing drugs from pharmacies or shops without a prescription, rather than visiting a healthcare facility (38). Similarly, in Ethiopia, 20% of patients with cough symptoms relied on antibiotics obtained from pharmacies or drug shops to treat these conditions (39).

Moreover, this tendency may also be driven by difficulties in accessing formal healthcare, where individuals face barriers in obtaining prescriptions from qualified healthcare providers (50). In some low- and middle-income countries (LMICs), the easy access to antibiotics, sometimes without proper regulation, exacerbates this issue (16). In China, 49.08% of patients with acute respiratory infections reported self-medicating (37), while in developing countries, 50% of patients with RTIs reportedly use leftover antibiotics to self-medicate (18).

Factors contributing to self-medication for RTIs in low- and middle-income regions include weak regulatory frameworks, the sharing of antibiotics, and the use of leftover medications from previous prescriptions, among others (51). Moreover, the high prevalence of RTIs, combined with the widespread availability of antibiotics, contributes to alarmingly high rates of self-medication, reaching up to 100% in some LMICs (52). While such practices are especially widespread in these regions, similar behaviors have also been observed in high-income countries such as those in Eastern Europe and Central Asia, where healthcare systems are more developed (30, 53). Collectively, these factors contributed to a 46% increase in global antibiotic consumption between 2000 and 2018 (30).

These factors underscore the broader challenges in managing respiratory illnesses in resource-limited settings, where both OTC use and self-medication without prescriptions are prevalent. Although antibiotics are considered prescription only according Kenya Pharmacy and Poisons Act (54), the use of over-the-counter (OTC) medications and self-treatment do not align with clinical guidelines. Consequently, such practices are often viewed as ‘irrational’ as they lead to the misuse of unprescribed antibiotics. This is a major obstacle to Antimicrobial Stewardship (AMS) in community settings in low- and middle-income countries (LMICs) (55), as it results in inappropriate treatment regimens and poor choices in drug selection and dosage.

As highlighted by (56), such practices can obscure or temporarily alleviate symptoms of infectious diseases, delay accurate diagnoses, and lead to inappropriate treatment with multiple course of antibiotics. This, in turn, increases both the likelihood of misuse and prolonged exposure to antibiotics which can select for resistant strains such as *Staphylococcus aureus Acinetobacter baumannii*, *Klebsiella pneumonia* and *Streptococcus pyogenes*, that may go on to colonize a host if conditions permit (57).

Beyond general patterns, this study found that gender and history of tuberculosis were the strongest predictors of antibiotic-seeking pathways. Women were significantly more likely than men to self-medicate, a finding that persisted even after adjustment. The gendered differences align with prior evidence showing that women are more likely than men to self-medicate hence a key determinant influencing access to antibiotics (15, 58–60). This is also echoed by (61), who, through a participatory arts-based study in Nepal, demonstrated the role of gender in shaping how people understand and respond to illness. For instance, women often lack access to formal healthcare and rely on over-the-counter antibiotics, sometimes using them inappropriately, contributing to the risk of antibiotic misuse and the development of resistance. Although data on women’s role in acquiring antibiotics in Kenya are limited, similar gender-related dynamics may influence antibiotic access and use in the Kenyan context. This pattern may be linked to women’s active role in family health and caregiving which may foster frequent decision-making regarding obtaining antibiotics, coupled with limited knowledge about appropriate antibiotic use, which together encourage self-medication.

Beyond gender, the study identified gross monthly income as a contextual factor associated with antibiotic-seeking behavior among respondents. Although this association may reflect confounded effects, it aligns with findings by (62), who reported that individuals with limited financial resources often resort to more affordable alternatives, such as self-medication, to obtaining antibiotics. Financial constraint helps explain why only about 26% of Kenyans are able to pay out-of-pocket for healthcare (63). Among those who do not seek formal healthcare, 43% self-medicate with non-prescription drugs, 20% cite cost as a barrier, and 10% report that their illness did not seem serious enough to justify the expense of medical care (62).

Conversely, a history of tuberculosis was associated with greater reliance on GP prescriptions and a significantly reduced likelihood of using alternative antibiotic-seeking pathway. Although such an exact finding has not been reported in the Kenyan context, evidence from Ethiopia show that 65% of patients with presumptive TB sought care from health facilities, compared to 17% who did not seek care at all and 18% who used alternative, mostly inappropriate, sources (64). Similarly, in Canada, patients with prior TB experience have been shown to have a greater healthcare facility use and continuity of formal care following treatment (65).

In addition to history of tuberculosis, the broader context of TB prevalence also shapes antibiotic-seeking behavior. In regions where TB is prevalent, individuals may be more accustomed to encountering antibiotics as part of routine healthcare interactions. For example, (66) describe how the use of trial-of-antibiotics to screen for TB among people with RTIs is common in Malawi, a practice also observed in many low- and middle-income countries. Such experiences can normalize empirical antibiotic use and reinforce the perception that antibiotics are necessary for treating respiratory symptoms, which may help explain the high rates of empirical antibiotic use observed in Kenya, where a majority (71.1%, 247 participants) reported using antibiotics without knowing the etiology of their coughs, compared to only 28.7% (100 participants) whose sputum samples were tested before antibiotic use in the current study.

While TB-related experiences appear to be a strong influence, other health-related conditions may also shape antibiotic-seeking patterns. For instance, high blood pressure was associated with antibiotic-seeking in the crude model, possibly reflecting repeated contact with healthcare systems and greater familiarity with medications. However, this may represent context-specific rather than consistent determinants of antibiotic use.

Beside the antibiotic-seeking pathways, this study revealed a concerning high rate of antibiotic consumption (86.7%, n=347) among patients with productive coughs in Nairobi County. Such elevated consumption aligns with prior studies from Kenya, where antibiotic use ranged from 70% to 87% for respiratory conditions (67). These figures reflect not only the local but also global trends where community-based antibiotic consumption ranges between 85% and 95% (68), representing a major risk factor for the rising problem of antibiotic resistance especially in resource-limited countries like Kenya (69).

According to (67), these rate could be reinforced by a widespread patient misconceptions, with over 66% to 74% believing antibiotics are effective against viral illnesses like colds and flu. Additionally, patients’ demand for or expectation of antibiotics, even in the absence of a prescription (70), contributes to the widespread misuse. Despite the fact that judicious use of antibiotics for bacteriologically confirmed RTIs can minimizes the emergence of antimicrobial resistance (71), the aforementioned patients factors reflects a broader pattern of overreliance on empirical use of antibiotics for respiratory illnesses, making this a high-risk population for the development and spread of antibiotic resistance.

Certainly, 71.1% (247) of participants took antibiotics without laboratory confirmation and only 28.7% (100) received tailored treatment based on sputum analysis, of whom 61% were diagnosed with tuberculosis, illustrating the missed opportunity for accurate diagnosis and targeted therapy. Similarly, in another study, 88% of patients suspected of having atypical Community Acquired Pneumonia (CAP) due to pathogens like Legionella, Mycoplasma, or Chlamydophila received empirical antibiotic treatment, despite not being bacteriologically infected as compared to the tailored treatment given to only 12% of patients who actually had CAP caused by these pathogens (72). Similarly, as highlighted by (73), the consuming of antibiotics without prior laboratory diagnosis is highly prevalent in Kenyan Hospitals, with 94% of prescriptions not based on laboratory data. Alarmingly, only 0.1% of cases involve the use of laboratory tools such as antibiotic susceptibility tests to treatment (73). This practice significantly increases the risk of unnecessary antibiotic exposure and the potential development ABR.

Empirical antibiotic therapy is exacerbated by structural limitation in Kenya healthcare systems including shortages of trained laboratory staff, inadequate supplies, and cost barriers to diagnostic testing (74), challenges that are across many African countries (75). This complicates the ability to distinguish between patients who genuinely need antibiotics and those seeking unnecessary prescriptions (76). Considering that only 10% of the respiratory illnesses required antibiotics (77), while many are viral in nature and self-limiting, widespread use of antibiotic empirically without addressing the root cause of the illness not only significantly contributes to the development of resistant bacterial pathogens but also lead to therapeutic failure of empirical treatment (78).

Additionally, patients’ behavior to often avoid care for respiratory illnesses until their symptoms worsen equally contributes to injudicious and irrational use of antibiotics (79). Avoiding formal care may partly stem from an overreliance on self-medication, which involves use of leftover antibiotics or shared medications to treat self-diagnosed respiratory illnesses, behaviors reported by 14.4% and 58.3% of participants in the study, respectively. Since self-medication does not align with clinical guidelines, this practice is considered irrational, as it often leads to poor drug selection and incorrect dosage, which can either obscure or temporarily alleviate symptoms (80), ultimately worsening outcomes in case of failed self-medication. These factors, combined with the absence of sufficient diagnostic support, perpetuate the cycle of unnecessary antibiotic use.

Use of antibiotics is further reinforced by individual-level factors, as revealed through regression analysis. In particular, participants living with HIV were significantly more likely to use antibiotics, possibly due to increased susceptibility to infections and routine prophylactic use (81). Despite smoking being a known risk factor for RTIs, study participants who reported a history of smoking and had productive cough were less likely to use antibiotics, a finding that could reflect a tendency to delay seeking formal healthcare, reduced perception of disease severity or patients differing perceptions of the necessity of treatment. Other significant predictors of antibiotic use included gender, age, history of tuberculosis, and high blood pressure, all of which may influence antibiotic seeking behaviors, risk perception, and overall interaction with the healthcare system.

These behavioral patterns may partially explain the frequent antibiotic use observed among participants in the study. Of the 347 participants, 233 (67.2%) reported multiple instances of antibiotic use, while only 32.9% (114) indicating occasional (single) use. This rate is substantially higher than figures reported in other regions. For instance, (82) found that only 10% of individuals RTIs received repeat antibiotic prescriptions within the same illness episode.

Notably, 14.4% (n=50) of participants had leftover antibiotics at home, and over half (52%) reused them. Similarly, a vast majority 58.3% of the 24 individuals who used shared antibiotics reported doing so repeatedly. These patterns reinforce the widespread misuse of antibiotics within the communities. Such informal, repeated use of antibiotics—outside the guidance of healthcare professionals may further contribute to poor treatment outcomes and resistance development. Moreover, the study revealed that these frequencies of antibiotic use are associated with income level, TB history, gender, and hypertension (p<0.05), suggesting that socioeconomic and health status jointly shape antibiotic use practices.

Beyond frequency, beta-lactams were most frequently used class of antibiotics among participants (66.0%, 229). Among them, amoxicillin and amoxicillin/clavulanic acid (amoxiclav) was particularly prevalent, reported by 90.8% (n=208) of those used beta-lactams. This suggests potential overuse and misuse of commonly accessible antibiotics. These findings align with other reports from Mozambique (83), which identified beta-lactams including amoxicillin and amoxiclav, as the most preferred antibiotics for self-diagnosed respiratory illness. These conditions often included sore throat, fever, throat pain, cough, common cold and influenza-like symptoms, all of which influence antibiotic use.

This high reliance on a single class of antibiotics mirrors concerns raised by (84), who noted the risks of resistance associated with overusing beta-lactams. Moreover, prior antibiotic exposures, particularly within the month preceding infection has been strongly associated with the development of resistant respiratory and urinary tract infections (85). Repeated antibiotic use has also been linked to increased risks of adverse effects, prolonged recovery, and escalating resistance rates (85, 86). Similarly, (87) found that even relatively low levels of frequent antibiotic use were associated with higher incidence of pneumonia. While some patients may seek repeated courses in the hope of a quick recovery, studies indicate that such practices offer minimal clinical benefit, particularly when antibiotics are used as precautionary rather than therapeutic agents.

A major strength of this study lies in its focus on the consumer side of antibiotic use, an area often overlooked in previous research. Unlike earlier studies that relied on secondary data, this research collected primary data directly from patients with productive coughs in Nairobi County. By exploring self-reported behaviors—such as preferred antibiotic sources and consumption patterns, the study provides a more realistic and nuanced understanding of the behavioral drivers of ABR. This patient-centered approach enhances the potential for designing actionable AMS interventions tailored to the needs of the patients.

However, this study has several limitations. Firstly, its cross-sectional design limits the ability to establish causal relationships between behaviors and antibiotic use outcomes. Second, reliance on self-reported data introduces potential recall and social desirability biases, which may affect the accuracy and reliability of the findings. Additionally, the limited use of laboratory diagnostics restricts the assessment of whether antibiotic use was clinically appropriate. This study focused solely on adult outpatients in an urban setting limiting generalizability to rural populations, children, or individuals presenting with different symptoms. Furthermore, the absence of longitudinal follow-up and qualitative data restricts the understanding of long-term outcomes and the underlying motivations for treatment choices. Lastly, while sociodemographic and health-related factors are briefly examined, the study did not deeply explore structural inequalities or cultural influences that may shape antibiotic-seeking pathways.

## Conclusions

The study highlights the high prevalence of antibiotic use among patients with productive coughs in Nairobi County, with multiple access sources including general practitioners, over-the-counter purchases, and self-medication. These patterns are influenced by sociodemographic factors such as gender, income level, and health history, particularly previous tuberculosis infection. A significant proportion of antibiotics were used empirically, without laboratory confirmation of the etiologic agents, with beta-lactams, especially amoxicillin and amoxicillin/clavulanic acid being the most frequently overused. These findings underscore a concerning trend of irrational antibiotic use that may accelerate antimicrobial resistance. As such enforcement of prescription-only regulations must be strengthened to curb unregulated access to antibiotics, while future research should evaluate community awareness and the effectiveness of stewardship interventions aimed at promoting rational antibiotic use.

## Acknowledgements

We acknowledge the University of Nairobi, Institute of Tropical and Infectious Diseases (UNITID), the Department of Medical Microbiology and Immunology, and the Kenya Medical Research Institute (KEMRI), Centre for Microbiology Research, for their institutional, logistical, laboratory, and technical support. We extend our sincere appreciation to the staff of the participating chest and tuberculosis clinics in Nairobi County for their administrative support, cooperation, and assistance during data collection. We are deeply grateful to all the patients who voluntarily participated in this study.

## Author’s contributions

A.K.M.: Conceptualization, methodology, data collection, data analysis, Original draft preparation and manuscript writing.

W.J.: Supervision, writing – review & editing, ethical oversight.

W.C.: Supervision, resources mobilization, writing – review & editing.

J.M.: Supervision, resources mobilization, technical support, writing – review & editing.

## Ethical approval

This study was approved by the Kenyatta National Hospital/University of Nairobi Ethics and Research Committee (Ref No. KNH-ERC/A/43). Written informed consent was obtained from all participants, and all data were handled confidentially

## Data Availability

Due to ethical restrictions related to participant confidentiality, the data supporting the findings of this study are available from the corresponding author upon reasonable request and with permission from the University of Nairobi Ethics/Kenyatta National Hospital Ethics Review Committee.

## Notes

### Competing Interest Statement

The authors have declared no competing interest.

### Funding Statement

The author(s) received no specific funding for this work.

### Author Declarations

KNH-UoN Ethics Research Committee (Ref No. KNH-ERC/A/43)

## References

1. World Health Organization. WHO Director-General’s opening remarks at the media briefing on COVID-192020 July 12 [cited 2025. Available from: https://www.who.int/director-general/speeches/detail/who-director-general-s-opening-remarks-at-the-media-briefing-on-covid-1911-march-2020.

2. Heikkinen T, Järvinen A. common cold. Lancet 2003;361(9351):51–9.

3. Song WJ, Chang YS, Faruqi S, Kim JY, Kang MG, Kim S, et al. The global epidemiology of chronic cough in adults: a systematic review and meta-analysis. The European Respiratory Journal. 2015;45(5):1479–81.

4. Chamberlain SA, Garrod R, Douiri A, Masefield S, Powell P, Bücher C, et al. The impact of chronic cough: a cross-sectional European survey. Lung. 2015;193(3):401–8.

5. Sharma S, Hashmi MF, Alhajjaj MS. Cough. In StatPearls. StatPearls Publishing: Treasure Island (FL); 2023. PMID: 33760535.

6. De Blasio F, Virchow JC, Polverino M, Zanasi A, Behrakis PK, Kilinç G, et al. Cough management: A practical approach. Cough. 2011;7(1):7.

7. Butler CC, Hood K, Verheij T, Little P, Melbye H, Nuttall J, et al. Variation in antibiotic prescribing and its impact on recovery in patients with acute cough in primary care: prospective study in 13 countries. BMJ. 2009;338:b2242.

8. Gulliford MC, Dregan A, Moore MV, Ashworth M, Staa TV, McCann G, et al. Continued high rates of antibiotic prescribing to adults with respiratory tract infection: survey of 568 UK general practices. BMJ Open. 2014;4(10):e006245.

9. Wang J, Wang P, Wang X, Zheng Y, Xiao Y. Use and prescription of antibiotics in primary health care settings in China. JAMA Internal Medicine. 2014;174(12):1914–20.

10. Muwanguzi TE, Yadesa TM, Agaba AG. Antibacterial prescription and the associated factors among outpatients diagnosed with respiratory tract infections in Mbarara Municipality, Uganda. BMC Pulmonary Medicine. 2021;21:374.

11. Nyamu N, Mbatia F, Van den Hombergh P, Jaarsma S, Agoi F, Shabani J, et al. Burden of upper respiratory tract infections in primary care facilities and excessive antimicrobial over-prescription: A community-oriented primary care project in rural Kenya. African Journal of Primary Health Care & Family Medicine. 2021;13(1):e1–e4.

12. Elnegaard S, Andersen RS, Pedersen AF. Self-reported symptoms and healthcare seeking in the general population – exploring “The Symptom Iceberg”. BMC Public Health. 2015;15:685.

13. Bennadi D. Self-medication: A current challenge. Journal of Basic and Clinical Pharmacy. 2013;5(1):19–23.

14. Cars O, Nordberg P. Antibiotic resistance – The faceless threat. International Journal of Risk & Safety in Medicine. 2005;17:103–10.

15. Torres NF, Chibi B, Kuupiel D. The use of non-prescribed antibiotics; prevalence estimates in low-and-middle-income countries. A systematic review and meta-analysis. Archives of Public Health. 2021;79:2.

16. Zhang T, Lambert H, Zhao L, Liu R, Shen X, Wang D, et al. Antibiotic Stewardship in Retail Pharmacies and the Access-Excess Challenge in China: A Policy Review. Antibiotics. 2022;11(2):141.

17. Godman B, Haque M, McKimm J, Abu Bakar M, Sneddon J, Wale J, et al. Ongoing strategies to improve the management of upper respiratory tract infections and reduce inappropriate antibiotic use particularly among lower and middle-income countries: Findings and implications for the future. Current Medical Research and Opinion. 2019;36(2):301–27.

18. Ocan M, Obuku EA, Bwanga F. Household antimicrobial self-medication: A systematic review and meta-analysis of the burden, risk factors and outcomes in developing countries. BMC Public Health. 2015;15:742.

19. Grigoryan L, Burgerhof JG, Degener JE, Deschepper R, Lundborg CS, Monnet DL, et al. Determinants of self-medication with antibiotics in Europe: the impact of beliefs, country wealth and the healthcare system. The Journal of Antimicrobial Chemotherapy. 2008;61(5):1172–9.

20. Muras M, Krajewski J, Nocun M, Godycki-Cwirko M. A survey of patient behaviours and beliefs regarding antibiotic self-medication for respiratory tract infections in Poland. Archives of Medical Science. 2013;9(5):854–7.

21. Hughes CM, McElnay JC, Fleming GF. Virus-induced secondary bacterial infection: a concise review. Therapeutics and Clinical Risk Management. 2001;24(14):1027–37.

22. Ruiz ME. Risks of self-medication practices. Current Drug Safety. 2010;5(4):315–23.

23. Bianco A, Papadopoli R, Mascaro V, Pileggi C, Pavia M. Antibiotic prescriptions to adults with acute respiratory tract infections by Italian general practitioners. Infection and Drug Resistance. 2018;11:2199–205.

24. Murray CJL, Ikuta KS, Sharara F, Moore CE. Global burden of bacterial antimicrobial resistance in 2019: a systematic analysis. The Lancet Planetary Health. 2022;399(10325):629–55.

25. Sulis G, Daniels B, Kwan A, Gandra S, Daftary A, Das J, et al. Antibiotic overuse in the primary health care setting: a secondary data analysis of standardised patient studies from India, China and Kenya. BMJ Global Health. 2020;5(9):e003393.

26. Saruti M, Brotherton J. Pattern of antibiotic prescription for upper respiratory tract infections among under-fives in outpatient clinics in Tharaka-Nithi County. Kabarak Journal of Research & Innovation. 2020;10(2):75–84.

27. Llor C, Hernández S, Bayona C, Moragas A, Sierra N, Hernández M, et al. A study of adherence to antibiotic treatment in ambulatory respiratory infections. International Journal of Infectious Diseases. 2013;17(3):e168–e72.

28. Hanson CL, Osberg M, Brown J, Durham G, Chin DP. Conducting patient-pathway analysis to inform programming of tuberculosis services: Methods. The Journal of Infectious Diseases. 2017;216(Suppl 7):S679–S85.

29. Prestinaci F, Pezzotti P, Pantosti A. Antimicrobial resistance: a global multifaceted phenomenon. Pathogens and Global Health. 2015;109(7):309–18.

30. Browne AJ, Chipeta MG, Haines-Woodhouse G, Kumaran EPA, Hamadani BHK, Zaraa S, et al. Global antibiotic consumption and usage in humans, 2000–18: a spatial modelling study. The Lancet Planetary Health. 2021;5(12):e893–e904.

31. Tola HH, Karimi M, Yekaninejad MS. Effects of sociodemographic characteristics and patients’ health beliefs on tuberculosis treatment adherence in Ethiopia: A structural equation modelling approach. Infectious Diseases of Poverty. 2017;6(1):167.

32. Kapatsa T, Lubanga AF, Bwanali AN, Harawa G, Mudenda S, Chipewa PC, et al. Behavioral and socio-economic determinants of antimicrobial resistance in Sub-Saharan Africa: A systematic review. Infection and Drug Resistance. 2025;18:855–73.

33. World Health Organization. People-centred approach to addressing antimicrobial resistance in human health: WHO core package of interventions to support national action plans. Geneva, Switzerland: 2023.

34. Guma SP, Godman B, Campbell SM, Mahomed O. Determinants of the Empiric Use of Antibiotics by general practitioners in South Africa: Observational, analytic, cross-sectional study. Antibiotics (Basel, Switzerland). 2022;11(10):1423.

35. Mukokinya MMA, Opanga S, Oluka M, Godman B. Dispensing of antimicrobials in Kenya: A cross-sectional pilot study and its implications. Journal of Research in Pharmacy Practice. 2018;7(2):77–82.

36. Finley CR, Chan DS, Garrison S, Korownyk C, Kolber MR, Campbell S, et al. What are the most common conditions in primary care? Systematic review. Canadian Family Physician Medecin de famille canadien. 2018;64(11):832–40.

37. Dai P, Qi L, Jia M, Li T, Ran H, Jiang M, et al. Healthcare-seeking behaviours of patients with acute respiratory infection: a cross-sectional survey in a rural area of southwest China. BMJ Open. 2024;14(2):e077224.

38. Emukule GO, Osoro E, Nyawanda BO, Ngere I, Macharia D, Bigogo G, et al. Healthcare-seeking behavior for respiratory illnesses in Kenya: implications for burden of disease estimation. BMC Public Health. 2023;23:353.

39. Senbeto M, Tadesse S, Tadesse T, Melesse T. Appropriate health-seeking behavior and associated factors among people who had cough for at least two weeks in northwest Ethiopia: A population-based cross-sectional study. BMC Public Health. 2013;13:1222.

40. Luis SF, Kamp N, Mitchell EM, Henriksen K, van Leth F. Health-seeking norms for tuberculosis symptoms in southern Angola: implications for behaviour change communications. The International Journal of Tuberculosis and Lung Disease: 2011;15(7):943–8.

41. Sánchez Choez X, Armijos Acurio ML, Jimbo Sotomayor RE. Appropriateness and adequacy of antibiotic prescription for upper respiratory tract infections in ambulatory health care centers in Ecuador. BMC Pharmacology & Toxicology. 2018;19(1):46.

42. Chandra Deb L, McGrath BM, Schlosser L, Hewitt A, Schweitzer C, Rotar J, et al. Antibiotic prescribing practices for upper respiratory tract infections among primary care providers: A descriptive study. Open Forum Infectious Diseases. 2022;9(7):ofac302.

43. Silverman M, Povitz M, Sontrop JM, Li L, Richard L, Cejic S, et al. Antibiotic prescribing for nonbacterial acute upper respiratory infections in elderly persons. Annals of Internal Medicine. 2017;166(11):765–74.

44. Fleming-Dutra KE, Hersh AL, Shapiro DJ, Bartoces M, Enns EA, File TM, Jr, et al. Prevalence of inappropriate antibiotic prescriptions among US ambulatory care visits, 2010–2011. JAMA Internal Medicine. 2016;315(17):1864–73.

45. Ayukekbong JA, Ntemgwa M, Atabe AN. The threat of antimicrobial resistance in developing countries: causes and control strategies. Antimicrobial Resistance & Infection Control. 2017;6:1–8.

46. Grijalva CG, Nuorti JP, Griffin MR. Antibiotic prescription rates for acute respiratory tract infections in US ambulatory settings. JAMA Internal Medicine. 2009;302(7):758–66.

47. Schroeck JL, Ruh CA, Sellick JA, Jr, Ott MC, Mattappallil A, Mergenhagen KA. Factors associated with antibiotic misuse in outpatient treatment for upper respiratory tract infections. Antimicrobial Agents and Chemotherapy. 2015;59(7):3848–52.

48. Belachew SA, Hall L, Selvey LA. Non-prescription dispensing of antibiotic agents among community drug retail outlets in Sub-Saharan African countries: A systematic review and meta-analysis. Antimicrobial Resistance and Infection Control. 2021;10:13.

49. Karambu I. Abuse of ‘Prescription Only’ Drugs on the Rise: Business Daily Africa; 2017 [August 25, 2017]. Available from: http://www.businessdailyafrica.com/Corporate-News/-/539550/1116580/-/vra01v/-/index.html.

50. Torres NF, Chibi B, Middleton LE, Solomon VP, Mashamba-Thompson TP. Evidence of factors influencing self-medication with antibiotics in low and middle-income countries: A systematic scoping review. Public Health. 2019;168:92–101.

51. Doan DA, Nguyen AD, Le GBTTXN, Phuong Lan Nguyen & Dai Xuan Dinh. Prevalence and associated factors of antibiotic self-medication and home storage among antibiotic users: a cross-sectional study in Vietnam. BMC Public Health. 2025;25:1940.

52. Sono TM, Yeika E, Cook A, Kalungia A, Opanga SA, Acolatse JEE, et al. Current rates of purchasing of antibiotics without a prescription across sub-Saharan Africa; rationale and potential programmes to reduce inappropriate dispensing and resistance. Expert Review of Anti-Infective Therapy. 2023;21(10):1025–55.

53. Grigoryan L, Haaijer-Ruskamp FM, Burgerhof JG, Mechtler R, Deschepper R, Tambic-Andrasevic A, et al. Self-medication with antimicrobial drugs in Europe. Emerging Infectious Diseases. 2006;12(3):452–9.

54. Government of Kenya. Pharmacy and Poisons Act [Internet]. Government of Kenya; 2012. p. 71.

55. Shamas N, Stokle E, Ashiru-Oredope D, Wesangula E. Challenges of implementing antimicrobial stewardship tools in Low to Middle Income Countries (LMICs). Infection Prevention in Practice. 2023;5(4):100315.

56. Satyanarayana S, Kwan A, Daniels B, Subbaraman R, McDowell A, Bergkvist S, et al. Use of standardised patients to assess antibiotic dispensing for tuberculosis by pharmacies in urban India: a cross-sectional study. The Lancet Infectious Diseases. 2016;16(11):1261–8.

57. Wesgate R, Evangelista C, Atkinson R, Shepard A, Adegoke O, Maillard JY. Understanding the risk of emerging bacterial resistance to over the counter antibiotics in topical sore throat medicines. Journal of Applied Microbiology. 2020;129(4):916–25.

58. El-Hawy RM, Ashmawy MI, Kamal MM, al. e. Studying the knowledge, attitude and practice of antibiotic misuse among Alexandria population. European Journal of Hospital Pharmacy. 2017;24(4):349–54.

59. Al-Tarawneh A, Ali T, Al-Taani GM. Public Patterns and Determinants of antibiotic self-Medication and antibiotic knowledge in southern Jordan. Antibiotics. 2024;13(1):98.

60. World Health Organization. Gender and health 2020 [cited 2025 July 12]. Available from: https://www.who.int/health-topics/gender.

61. Jones N, Mitchell J, Cooke P, Baral S, Arjyal A, Shrestha A, et al. Gender and antimicrobial resistance: What can we learn from applying a gendered lens to data analysis using a participatory arts case study? Frontiers in Global Women’s Health. 2022;3:745862.

62. Ngugi AK, Agoi F, Mahoney MR, Lakhani A, Mang’ong’o D, Nderitu E, et al. Utilization of health services in a resource-limited rural area in Kenya: Prevalence and associated household-level factors. PLoS One. 2017;12(2):e0172728.

63. Barasa E, Nguhiu P, McIntyre D. Measuring progress towards Sustainable Development Goal 3.8 on universal health coverage in Kenya. BMJ Global Health. 2018;3(3):e000904.

64. Gamtesa DF, Tola HH, Mehamed Z, al. e. Health care seeking behavior among presumptive tuberculosis patients in Ethiopia: a systematic review and meta-analysis. BMC Health Services Research. 2020;20:445.

65. Romanowski K, Karim ME, Gilbert M, Cook VJ, Johnston JC. Distinct healthcare utilization profiles of high healthcare use tuberculosis survivors: A latent class analysis. PLoS One. 2023;18(9):e0291997.

66. Divala TH, Fielding KL, Sloan DJ, French N, Nliwasa M, MacPherson P, et al. Accuracy and consequences of using trial-of-antibiotics for TB diagnosis (ACT-TB study): Protocol for a randomised controlled clinical trial. BMJ Open. 2020;10(3):e033999.

67. Omulo S, Thumbi SM, Lockwood S, Verani JR, Bigogo G, Masyongo G, et al. Evidence of superficial knowledge regarding antibiotics and their use: Results of two cross-sectional surveys in an urban informal settlement in Kenya. PLoS One. 2017;12(10):e0185827.

68. Duffy E, Ritchie S, Metcalfe S, Van Bakel B, Thomas MG. Antibacterials dispensed in the community comprise 85%–95% of total human antibacterial consumption. Journal of Clinical Pharmacy and Therapeutics. 2018;43(1):59–64.

69. Bebell LM, Muiru AN. Antibiotic use and emerging resistance: How can resource-limited countries turn the tide? Global Heart. 2014;9(3):347–58.

70. Almeshal N, Foot H, Clarke AL, Chan AHY, Horne R. Understanding patient demand for and use of antibiotics for upper respiratory tract infection: A qualitative application of the Necessity-Concerns Framework in Saudi Arabia. Frontiers in Pharmacology. 2024;15:1399698.

71. Regasa B. Drug resistance patterns of bacterial pathogens from adult patients with pneumonia in Arba Minch Hospital, South Ethiopia. Journal of Medical Microbiology and Diagnosis. 2014;3(4):151–4.

72. Lorentzen MH, Rosenvinge FS, Lassen AT, Graumann O, Laursen CB, Mogensen CB, et al. Empirical antibiotic treatment for community-acquired pneumonia and accuracy for Legionella pneumophila, Mycoplasma pneumoniae, and Chlamydophila pneumoniae: A descriptive cross-sectional study of adult patients in the emergency department. BMC Infectious Diseases. 2023;23(1):580.

73. Maina M, Mwaniki P, Odira E, Kiko N, McKnight J, Schultsz C, et al. Antibiotic use in Kenyan public hospitals: Prevalence, appropriateness and link to guideline availability. International Journal of Infectious Diseases. 2020;99:10–8.

74. Dat VQ, Dat TT, Hieu VQ, Giang KB, Otsu S. Antibiotic use for empirical therapy in the critical care units in primary and secondary hospitals in Vietnam: a multicenter cross-sectional study. The Lancet Regional Health – Western Pacific. 2021;18:100306.

75. Talaat M, Saied T, Kandeel A, Abo El-Ata GA, El-Kholy A, Hafez S, et al. A point prevalence survey of antibiotic use in 18 hospitals in Egypt. Antibiotics. 2014;3(3):450–60.

76. Istúriz RE, Carbon C. Antibiotic use in developing countries. Infection Control & Hospital Epidemiology. 2000;21(6):394–7.

77. Pouwels KB, Dolk FCK, Smith DRM, Robotham JV, Smieszek T. Actual versus ’ideal’ antibiotic prescribing for common conditions in English primary care. The Journal of Antimicrobial Chemotherapy. 2018;73(suppl_2):19–26.

78. Khan S, Priti S, Ankit S. Bacteria etiological agents causing lower respiratory tract infections and their resistance patterns. Iranian Biomedical Journal. 2015;19(4):240–6.

79. Burton DC, Flannery B, Onyango B, Larson C, Alaii J, Zhang X, et al. Healthcare-seeking behaviour for common infectious disease-related illnesses in rural Kenya: a community-based house-to-house survey. Journal of Health, Population, and Nutrition. 2011;29(1):61–70.

80. Satyanarayana S, Kwan A, Daniels B, Subbaraman R, McDowell A, Bergkvist S, et al. Use of standardised patients to assess antibiotic dispensing for tuberculosis by pharmacies in urban India: a cross-sectional study. The Lancet Infectious Diseases. 2016;16(11):1261–8.

81. Faiela C, Sevene E. Antibiotic prescription for HIV-positive patients in primary health care in Mozambique: A cross-sectional study. Southern African Journal of Infectious Diseases. 2022;37(1):340.

82. Lalmohamed A, Venekamp RP, Bolhuis A, Souverein PC, van de Wijgert JHHM, Gulliford MC, et al. Within-episode repeat antibiotic prescriptions in patients with respiratory tract infections: A population-based cohort study. The Journal of Infection. 2024;88(4):106135.

83. Torres NF, Solomon VP, Middleton LE. Identifying the commonly used antibiotics for self-medication in urban Mozambique: A qualitative study. BMJ Open. 2020;10:e041323.

84. Machado-Duque ME, García DA, Emura-Velez MH, Gaviria-Mendoza A, Giraldo-Giraldo C, Machado-Alba JE. Antibiotic prescriptions for respiratory tract viral infections in the Colombian population. Antibiotics. 2021;10(7):864.

85. Costelloe C, Metcalfe C, Lovering A, Mant D, Hay AD. Effect of antibiotic prescribing in primary care on antimicrobial resistance in individual patients: systematic review and meta-analysis. BMJ. 2010;340:c2096.

86. van Staa TP, Palin V, Li Y, Welfare W, Felton TW, Dark P, et al. The effectiveness of frequent antibiotic use in reducing the risk of infection-related hospital admissions: results from two large population-based cohorts. BMC Medicine. 2020;18(1):40.

87. Gulliford MC, Moore MV, Little P, Hay AD, Fox R, Prevost AT, et al. Safety of reduced antibiotic prescribing for self limiting respiratory tract infections in primary care: cohort study using electronic health records. BMJ. 2016;354: i3410.

